# Evaluating Deep Learning–Based Quantification of Breast Arterial Calcification on Mammography for Cardiovascular Risk Assessment

**DOI:** 10.64898/2026.06.16.26355800

**Authors:** Pranav Singh, Samantha Platt, Olivia Bussey, Laura Heacock, Antonio Verdone, Weixi Chen, Harmony R. Reynolds, Chang Yu, Yiqiu Shen, Miriam A. Bredella

**Author notes:** Corresponding author: Miriam A. Bredella, MD, MBA, Department of Radiology, NYU Langone Health and NYU Grossman School of Medicine, Translational Research Building 743, 227 East 30th Street, New York, NY 10016. Funding information: None. **Data Sharing:** All data generated or analyzed during the study are included in the published paper.

## Abstract

**Purpose:** To develop and evaluate a deep learning model for automated quantification of breast arterial calcification (BAC) on screening mammography and to assess whether AI-derived BAC burden predicts major adverse cardiovascular events (MACE) in women.

**Methods:** In this retrospective study, 202,006 women who underwent screening mammography without history of MACE were included. A BAC segmentation model was trained on an expert-annotated dataset using a multi-task U-Net with a ResNet-18 encoder to detect and segment BAC. BAC burden was quantified as area (mm²) from model-generated masks using DICOM pixel spacing and categorized by tertiles into low, intermediate, and high. The PREVENT score and incident MACE were identified from electronic health records. Cox proportional hazards models were developed to evaluate AI-derived BAC burden and PREVENT score alone, and combined models for 5- and 10-year cardiovascular risk prediction.

**Results:** Among 202,006 women (mean age 54.8±11.7 years), 23.1% had AI-detected BAC, and 7,701 (3.8%) developed incident MACE during a median follow-up of 7.5 years. On the geographically held-out test set, the BAC model achieved an AUROC of 0.97, Dice score of 0.6678, and Pearson correlation of 0.961 between AI-derived and manually annotated BAC burden. BAC burden increased with age and was higher among women who developed MACE. Five-year MACE incidence increased across BAC categories from 1.5% in women without BAC to 6.9% in those with high BAC burden. BAC burden alone showed modest prediction of MACE, with 5-year and 10-year AUROCs of 0.661 and 0.650, respectively, while PREVENT achieved AUROCs of 0.781 and 0.771. Adding BAC to PREVENT produced minimal improvement in discrimination.

**Conclusion:** Deep learning-based BAC quantification from routine mammography is feasible, accurate, and associated with future cardiovascular risk. Although BAC added little to PREVENT for overall discrimination, it may serve as a scalable opportunistic imaging biomarker to identify women at elevated cardiovascular risk and support preventive care.

## Introduction

Cardiovascular disease is the leading cause of death among women worldwide, despite increasing attention to primary prevention and treatment ^1,2^. Many women, particularly younger and middle-aged women, are often misclassified as “low” or “intermediate risk” by traditional prediction tools such as the Framingham Heart Score, which was derived from male-dominant cohorts ^3^. This underestimation of risk contributes to delayed diagnosis, undertreatment ^4–6^, and preventable morbidity and mortality, underscoring the need for more sex-specific approaches to cardiovascular risk assessment.

Arterial calcifications are major risk factors for cardiovascular disease events such as heart attack and stroke^7,8^ and typically affect blood vessels throughout the body. Arterial calcifications can be detected via medical imaging before cardiovascular events occur, presenting an opportunity for early intervention ^8,9^. However, this requires specialized testing that must be ordered by a clinician who is already considering increased cardiovascular risk for that patient, limiting early detection of asymptomatic patients.

“Opportunistic imaging”, which involves the systematic quantification of imaging biomarkers (e.g., arterial calcification) through imaging tests already performed for other clinical reasons, offers a powerful opportunity for early risk detection ^10^. Specifically, arterial calcification can be detected on screening mammograms, which are performed in 40 million women annually in the U.S, and these breast arterial calcifications (BAC) are associated with increased future cardiovascular disease event rates ^11–16^. Despite this compelling evidence, BAC are rarely quantified or systematically reported on routine mammograms ^17,18^.

Recent advances in artificial intelligence (AI) and deep learning allow the automated quantification of vascular calcification ^19^. Leveraging mammograms to automatically quantify BAC could enable early identification of women at risk for cardiovascular events, even before clinical suspicion arises ^18^. The purpose of our study was to develop an automated deep learning algorithm to quantify BAC on screening mammograms and to investigate the value of deep learning-enabled BAC quantification as a predictor of cardiovascular events.

## Methods

This retrospective study was approved by the NYU Grossman School of Medicine institutional review board and was compliant with the Health Insurance Portability and Accountability Act. The requirement for informed consent was waived. The study was conducted in compliance with the TRIPOD reporting guidelines ^20^.

### Patient Population

We identified 211,034 women aged 18 years or older who underwent full-field digital mammography (FFDM) screening as part of routine clinical care at NYU Langone Health between January 2010 and December 2022. The index mammogram was defined as the first eligible screening mammogram during the study period. We excluded 9,028 patients with a documented major adverse cardiovascular event (MACE) before the index mammogram, resulting in a final study cohort of 202,006 patients. There were no further exclusion criteria.

### Data Collection

Mammographic images and electronic health record (EHR) data were obtained for all included patients. Mammographic data consisted of standard screening FFDM examinations, including four views: right and left craniocaudal and right and left mediolateral oblique views. FFDM examinations were acquired across multiple vendors, including Hologic, Siemens, and GE Healthcare systems, and raw Digital Imaging and Communications in Medicine (DICOM) files were retrieved. Clinical variables were extracted from the EHR (Epic Systems, Verona, WI, United States) for cardiovascular risk modeling and calculation of the Predicting Risk of Cardiovascular Disease EVENTs (PREVENT) score ^21^. Extracted variables included age, sex, race, ethnicity, body mass index, systolic and diastolic blood pressure, high-density lipoprotein cholesterol, low-density lipoprotein cholesterol, total cholesterol, estimated glomerular filtration rate, glycated hemoglobin, and smoking status. For each patient, clinical variables were temporally mapped to the mammogram, with the closest available measurement on or before the mammogram date used for analysis.

### BAC Dataset Construction

We constructed an expert-annotated dataset of 2,676 FFDM images for BAC model development and validation. Images were randomly selected using an enrichment strategy designed to balance BAC-positive and BAC-negative cases for model training and validation. Candidate images were preselected based on radiology report text, including screening examinations with no mention of calcifications and examinations with explicit mention of calcifications. This report-based approach was used only for sampling; final BAC labels were determined by manual image review and annotation. A radiology resident (S.P.) manually annotated BAC using a HIPAA-compliant, intranet-based annotation platform deployed in our Picture Archiving and Communication System (PACS, Visage Imaging, Inc., San Diego, CA, United States). BAC was delineated with a freehand drawing tool when present, and images without visible BAC were assigned empty masks. For quality control, an attending breast radiologist (L.H.) with nine years of post-fellowship experience reviewed the annotations. This resulted in a dataset with 776 BAC-positive images and 1900 BAC-negative images. All images from the same patient were assigned to a single split to prevent patient-level data leakage. The dataset was divided into training, validation, and test sets containing 1907, 360, and 409 images, respectively.

### BAC Segmentation Model Development

A deep learning pipeline was developed to generate BAC segmentation maps from screening FFDM images. Before model training, each image was preprocessed by cropping to a standardized resolution of 2,944 × 1,920 pixels while preserving the breast region of interest. The complete image preprocessing pipeline has been described in detail in prior work ^22^. The BAC segmentation model was trained on individual FFDM images using a multi-task convolutional neural network. The model used a U-Net architecture ^23^ with a ResNet-18 ^24^. Convolutional Neural Network initialized from scratch as an encoder and an additional image-level classification head. The network jointly generated two outputs: a pixel-level BAC segmentation map and an image-level probability of BAC presence. The segmentation task was optimized using Focal-Tversky loss ^25^, and the image-level classification task was optimized using binary cross-entropy loss. The final training objective was a weighted combination of the classification and segmentation losses as described in the following equation:

loss = (1 − λ) • loss_segementation + λ loss_classification. We swept across various λ [0.1, 0.3, 0.5 and 0.7] to finally select λ = 0.1 as the most suitable balance. The model was trained for 50 epochs with early stopping based on validation performance using a patience of three epochs. Training used Adam optimizer, an initial learning rate of 1.135e-05, batch size of 4, and image augmentation including resizing all images to 2560 x 1664, channel modifier to increase number of channels from one to three and finally channel normalization. The final model was selected based on least loss on the validation set. During inference, the trained model generated BAC probability scores and binary BAC segmentation masks, which were used for downstream BAC quantification.

### BAC Quantification

Following segmentation, BAC burden was quantified from the binary BAC mask generated by the trained model. For each FFDM image, the model output probability map was converted to a binary mask using a prespecified threshold of 0.5. The number of foreground pixels within the binary BAC mask was then summed to obtain the image-level BAC pixel count. To convert pixel count into calibrated physical area, the DICOM-encoded Pixel Spacing attribute (tag 0028,0030) was extracted for each image. Pixel spacing represents the center-to-center distance between adjacent pixels in millimeters. The physical BAC area for each image was calculated as the number of foreground BAC pixels multiplied by the product of the row and column pixel spacing values, yielding BAC burden in mm². For each screening examination, exam-level BAC burden was calculated by averaging the BAC area across all FFDM views.

### Cardiovascular Outcome Definitions

The primary outcome was incident major adverse cardiovascular events (MACE) after the index screening mammogram. Patients with a documented MACE before the index mammogram were excluded. For patients with incident MACE, event time was defined as the date of the first qualifying diagnosis or procedure code after the index mammogram. Patients without MACE were censored at the last available in-person clinical encounter.

Cardiovascular outcomes were identified from the EHR using ICD-9/10 diagnosis codes and procedure codes. Cardiovascular death was not included because cause-specific mortality could not be reliably determined from the EHR ^26^. MACE was defined in consultation with a cardiologist (HRR) and included acute myocardial infarction (I21-22), ischemic or unspecified stroke (I63-64), hemorrhagic stroke (I60-62), unstable angina or other acute ischemic heart disease (I20.0 or I24 other than I24.1, I25.110), cardiac arrest (I46.2), and acute heart failure. Acute myocardial infarction was identified using ICD-10 codes beginning with I21 or I22 (I50, excluding chronic heart failure codes I50.22, I50.32, and I50.42).

### Cardiovascular Risk Cohort

For cardiovascular risk prediction modeling, we excluded patients whose mammograms were used for BAC segmentation model development to avoid overlap between BAC model development and downstream cardiovascular risk evaluation. After this exclusion, the risk modeling cohort included 202,006 patients, of whom 7701 experienced incident MACE during follow-up (3.81%). The remaining cohort was split by geographic location to evaluate model generalizability across patient populations. Patients from Manhattan, Queens, Staten Island, and Long Island were used for model development and were further divided into training, validation, and internal test sets comprising 114,292, 19,048, and 57,147 patients, respectively. The training, validation, and internal test sets were generated at the patient level, with all mammographic examinations from the same patient assigned to the same split. The proportion of patients with MACE in the development cohort was 3.57%. Patients from Brooklyn were reserved as a geographically held-out test cohort to assess model performance in a distinct patient population. This held-out cohort included 11,519 patients, of whom 539 experienced MACE during follow-up (4.68%). Baseline demographic and clinical characteristics of the development and geographically held-out cohorts are summarized in **Table 1**.

**Table 1:** Characteristics of the Development Cohort, Internal Test Cohort, and Geographically Held-Out Brooklyn Test Cohort.

| Characteristic | Development cohort | Internal test cohort | Brooklyn held-out test cohort |
| --- | --- | --- | --- |
| **Patients, No.** | 133,340 | 57,147 | 11,519 |
| **Age, years** | 54.0 [45.0–63.0] | 54.0 [45.0–64.0] | 51.0 [44.0–60.0] |
| **Follow-up duration, years** | 7.80 [5.29, 9.88] | 7.81 [5.33, 9.88] | 6.09 [4.37, 7.85] |
| **Screening mammograms, No.** | 533,360 | 228,588 | 46,076 |
| **Screening mammograms per patient** | 4 | 4 | 4 |
| **Race** |  |  |  |
| White | 80246 (60.2%) | 34576 (60.5%) | 3734 (32.4%) |
| Black or African American | 15737 (11.8%) | 10352 (18.1%) | 1770 (15.4%) |
| Asian | 8483 (6.4%) | 6753 (11.8%) | 541 (4.7%) |
| Other/Unknown | 24653 (18.5%) | 3641 (6.4%) | 1614 (14.0%) |
| Hispanic or Latino | 2826 (2.1%) | 1227 (2.1%) | 3231 (28.0%) |
| Multiracial/Multiple Races | 1007 (0.8%) | 398 (0.7%) | 155 (1.3%) |
| American Indian or Alaska Native | 162 (0.1%) | 91 (0.2%) | 242 (2.1%) |
| Native Hawaiian or Pacific Islander | 159 (0.1%) | 75 (0.1%) | 191 (1.7%) |
| Middle Eastern or North African | 67 (0.1%) | 34 (0.1%) | 41 (0.4%) |
| **Ethnicity** |  |  |  |
| White | 80246 (60.2) | 34576 (60.5) | 3734 (32.4) |
| Black or African American | 15737 (11.8) | 10352 (18.1) | 1770 (15.4) |
| Asian | 8483 (6.4) | 6753 (11.8) | 541 (4.7) |
| Hispanic | 2826 (2.1) | 1227 (2.1) | 3231 (28) |
| Other | 25041 (18.8) | 3841 (6.7) | 2088(18.1) |
|  | 1007 (0.8) | 398 (0.7) | 155 (1.3) |
| **BMI, kg/m<sup>2</sup>** | 26.3 [22.5–31.0] | 26.3 [22.5–30.9] | 28.3 [24.2–32.7] |
| **Systolic blood pressure, mm Hg** | 123.0 [113.0–134.0] | 123.0 [112.0–134.0] | 124.0 [114.0–136.0] |
| **Diastolic blood pressure, mm Hg** | 75.0 [70.0–80.0] | 75.0 [70.0–80.0] | 76.0 [70.0–82.0] |
| **Total cholesterol, mg/dL** | 195.0 [170.0–222.0] | 194.0 [170.0–221.0] | 190.0 [166.0–217.0] |
| **HDL cholesterol, mg/dL** | 62.0 [52.0–75.0] | 62.0 [52.0–75.0] | 55.0 [46.0–66.0] |
| **LDL cholesterol, mg/dL** | 109.0 [87.0–132.0] | 108.0 [87.0–131.0] | 107.0 [86.0–130.0] |
| **eGFR, mL/min/1.73 m<sup>2</sup>** | 92.0 [77.0–104.9] | 92.0 [77.0–104.0] | 100.0 [85.0–113.0] |
| **HbA1c, %** | 5.5 [5.3–5.9] | 5.5 [5.3–5.9] | 5.7 [5.4–6.1] |
| **Current or prior tobacco use** | 31440 (23.5%) | 13447 (23.5%) | 2362 (20.5%) |
| **10-year PREVENT score, %** | 2.9 [1.2–7.0] | 2.9 [1.2–7.0] | 2.8 [1.2–7.2] |
| **AI-derived BAC presence** | 30614 (23.0%) | 13153 (23.0%) | 2911 (25.3%) |
| **BAC burden among BAC-positive patients, mm<sup>2</sup>** | 1.4 [0.0–7.8] | 1.4 [0.0–7.9] | 2.1 [0.0–12.2] |
| **Incident MACE** | 5057 (3.8%) | 2105 (3.7%) | 539 (4.7%) |

### Cardiovascular Risk Modeling

We developed a survival analysis pipeline to predict the first onset of MACE within a 5-and 10-year cumulative horizon using one or more combinations of biomarkers. For each patient with MACE, the event date was assigned as the earliest confirmed MACE date. MACE-negative patients were censored at the date of last healthcare contact. Follow-up time was computed as the difference between the index study date and the event/censoring date. Patients with missing follow-up times were excluded. Follow-up was administratively censored at 10 years for patients’ event-free beyond that threshold.

Four Cox Proportional Hazards (Cox-PH) models were developed: (i) AI-derived BAC burden (mm²) alone; (ii) the ACC/AHA PREVENT 10-year cardiovascular disease risk score alone; (iii) a combined model incorporating both BAC area (mm²) with the ACC/AHA PREVENT score and (iv) a combined model incorporating standard cardiovascular risk factors (age, sex, BMI, serum lipids, blood pressure, glomerular filtration rate, glycated hemoglobin) and AI-derived BAC burden (mm²). For each of them, hyperparameters were selected using Bayesian optimization via Optuna (version ≥ 3.0) with the Tree-structured Parzen Estimator (TPE) sampler. The search was conducted over penalizer in [1×10⁻D, 5.0] search space on log-uniform scale and on l1 ratio in [0,1] uniform search space. Each trial fit the Cox model on the training set and evaluated the concordance index (C-index) on the validation set. A maximum of 50 trials was run per horizon, and the parameter combination maximizing validation C-index was selected. Post-hoc Platt scaling was applied for calibration. The calibration method was fit exclusively on the validation set after Cox model training was complete. No recalibration was performed on the test set.

To evaluate the predictive value of BAC burden, we modeled BAC area as a continuous quantitative measure derived from automated image analysis. Because BAC area was right-skewed (**Figure 1**) and included zero values, we applied a log(1+x) transformation before modeling. This transformation accommodates zero values without adding an arbitrary constant and reduces the influence of extreme values.

**Figure 1.**
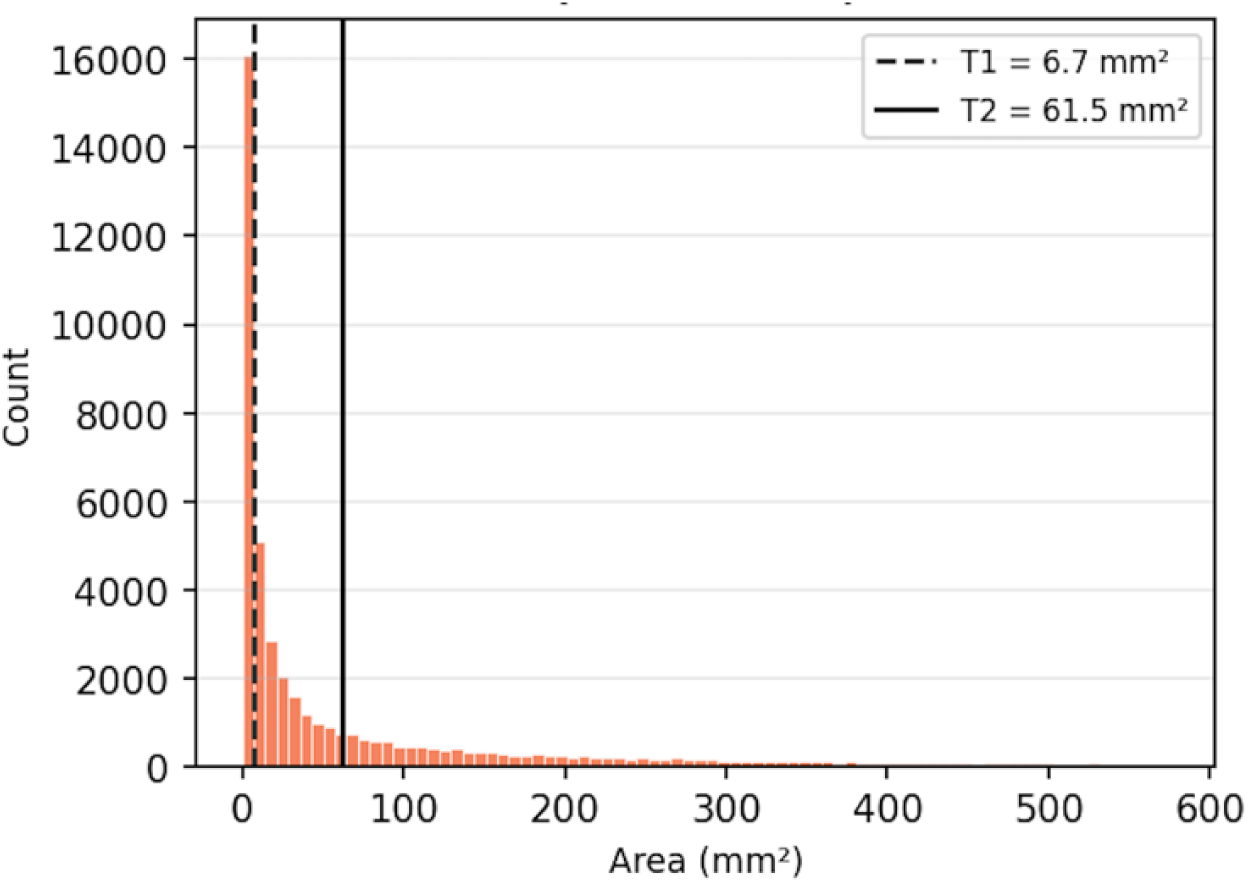
Right-skewed distribution of BAC burden among BAC-positive women. Raw breast arterial calcification (BAC) area (mm²) among BAC-positive patients (n = 46,678), truncated at the 95th percentile (574.7 mm²) to aid visualization of the bulk distribution. The marked positive skew reflects that most women with detectable BAC carry a modest calcification burden, while a minority exhibit extensive vascular calcification. Vertical lines denote tertile cut points with first tertile (T1) at 6.7 mm² and second tertile (T2) at 61.5 mm².

### PREVENT Score

Clinical variables were extracted from the EHR to calculate the 10-year PREVENT score ^21^. We used the American Heart Association PREVENT equations designed to estimate 10-year total cardiovascular disease risk among adults without known cardiovascular disease. Because this study focused on near- to intermediate-term cardiovascular risk after screening mammography, we used the 10-year PREVENT estimate rather than the 30-year estimate. For each patient, PREVENT input variables were selected as the closest available measurements to the index mammogram date. Variables included total cholesterol, HDL cholesterol, systolic blood pressure, sex, estimated glomerular filtration rate, age, diabetes status, current tobacco use, and use of antihypertensive or statin therapy. PREVENT was evaluated as a clinical benchmark for cardiovascular risk prediction. Missing clinical variables were handled using multiple imputations by chained equations. Three imputed datasets were generated, each using five iterations and a different random seed to reflect imputation uncertainty. PREVENT scores were calculated separately within each imputed dataset, and the final analyses used the third imputed dataset. The resulting PREVENT score, expressed as a continuous covariate into Cox Proportional Hazards models for 5- and 10-year MACE prediction.

### Risk Stratification of AI-derived BAC Burden

Patients were stratified according to AI-derived BAC burden into four categories: no BAC, low BAC burden, intermediate BAC burden, and high BAC burden. Among patients with BAC, low, intermediate, and high BAC burden were defined by tertiles of AI-derived BAC area in the internal test cohort, corresponding to <33.3%, 33.3% to 66.6%, and >66.6% of the BAC-positive distribution, respectively. These thresholds were then applied to the study cohorts for downstream analyses. Five-year and 10-year MACE incidence rates were calculated across BAC burden categories. Differences in event incidence across BAC burden categories were assessed using chi-square test. Time-to-event analyses were performed using Kaplan-Meier curves, and differences in MACE-free survival across BAC burden categories were assessed using the log-rank test.

### Statistical Analysis

Baseline characteristics were summarized for the overall cohort and by data split. Continuous variables were reported as median with interquartile range, and categorical variables as counts and percentages. Between-cohort differences were assessed using Wilcoxon rank-sum tests for continuous variables and chi-square or Fisher exact tests for categorical variables, as appropriate.

The technical performance of the BAC segmentation model was evaluated on the held-out annotated test set. BAC detection was assessed using Area Under the Receiver Operating Characteristic (AUROC). Segmentation performance was assessed using Dice similarity coefficient and intersection-over-union. BAC area quantification was assessed using mean absolute error, normalized mean absolute error, Pearson correlation coefficient, and R². Because BAC area was right-skewed, BAC burden was log-transformed for regression analyses.

Cardiovascular risk prediction was evaluated for MACE across 5-year and 10-year horizons. Models included PREVENT score alone, AI-derived BAC burden alone, standard cardiovascular clinical risk factors alone, standard cardiovascular clinical risk factors plus BAC burden, and PREVENT score plus BAC burden. Models were trained on the development cohort and evaluated on the internal test and geographically held-out test cohorts. Discrimination was assessed using AUROC for fixed-horizon prediction and Harrell’s C-index for time-to-event analysis, with 95% confidence intervals estimated using 1,000 stratified bootstrap iterations. Pairwise AUROC comparisons were performed using DeLong’s test, and C-index differences were compared using bootstrap testing. Calibration was assessed using calibration plots, calibration slope and intercept, and Brier score. Subgroup analyses were performed by age group, race, ethnicity, BAC presence, and PREVENT risk category. All statistical tests were two-sided, and p<0.05 was considered statistically significant. Analyses were performed using Python 3.10 with pandas, numpy, scipy, statsmodels, scikit-learn, lifelines, matplotlib, and seaborn packages.

## Results

### Study Population

The final cohort included 202,006 women, with a mean age of 54.8 ± 11.7 years (median 54.0 [45.0–63.0] years) and a median follow-up of 7.5 [5.0–9.7] years. During follow-up, 7,701 (3.8%) women developed incident MACE. At 5 years, the MACE event rate was 2.2% (4,405 events); at 10 years, 3.6% (7,278 events). AI-detected BAC was present in 23.1% of the overall cohort (46,678 women), with a median BAC burden of 19.51 [3.66–111.37] mm² among BAC-positive patients. The development cohort included 133,340 women (MACE event rate: 3.8%). The internal test cohort included 57,147 women (MACE event rate 3.7%). The geographically held-out Brooklyn test cohort included 11,519 women (MACE event rate 4.7%). Compared with the internal test cohort, the Brooklyn cohort differed in demographic and clinical characteristics (**Table 1**).

### BAC Quantification Performance

In the held-out annotated test set of 2,676 FFDM images, the BAC model achieved an image-level BAC detection AUROC of 0.97. For pixel-level BAC segmentation, the model achieved a Dice score of 0.6678. For BAC burden quantification (**Figure 2**), AI-derived BAC burden had a Pearson correlation coefficient of 0.961 with manually annotated BAC burden. The mean absolute error was 21.5 mm², and the root mean squared error was 52.9 mm². The mean bias was +10.3 mm², indicating a slight systematic overestimation of BAC burden by the AI model relative to manual annotation. Among BAC-positive images, the Pearson correlation coefficient was 0.950, and the mean absolute error was 58.5 mm². Among BAC-negative images, the mean absolute error was 7.2 mm², reflecting cases where model predicted a small but nonzero BAC area despite the absence of manually annotated BAC. **Figure 3** shows representative examples of AI-derived BAC segmentation, including two cases with close agreement between manual annotation and model output and one failure case with incomplete or false-positive segmentation.

**Figure 2.**
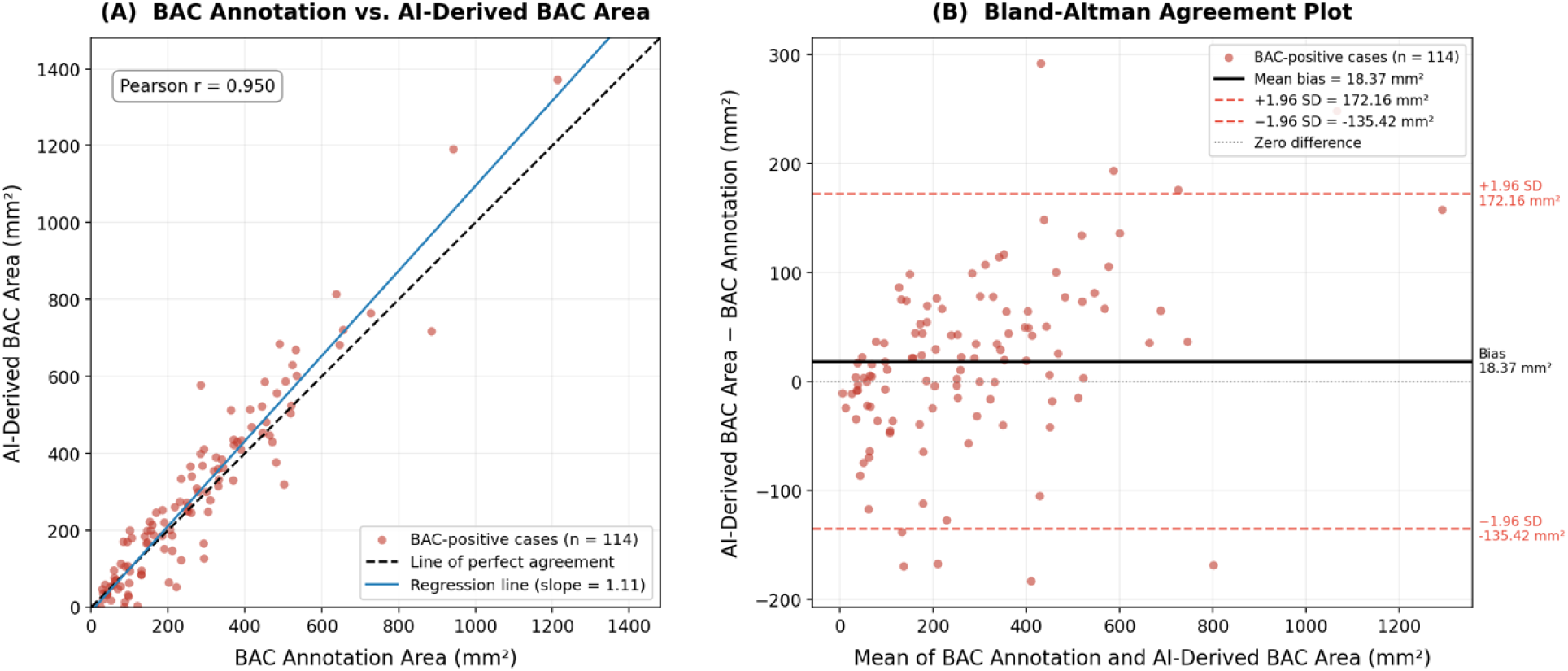
Agreement between manual BAC annotation and AI-derived BAC area. (A) Scatter plot comparing manually annotated breast arterial calcification (BAC) area with AI-derived BAC area among BAC-positive images in the held-out annotated test set. The dashed line indicates perfect agreement, and the solid line indicates the fitted regression line. (B) Bland–Altman plot showing the difference between AI-derived and manually annotated BAC area against their mean. The solid line indicates the mean bias, and dashed lines indicate the 95% limits of agreement.

**Figure 3.**
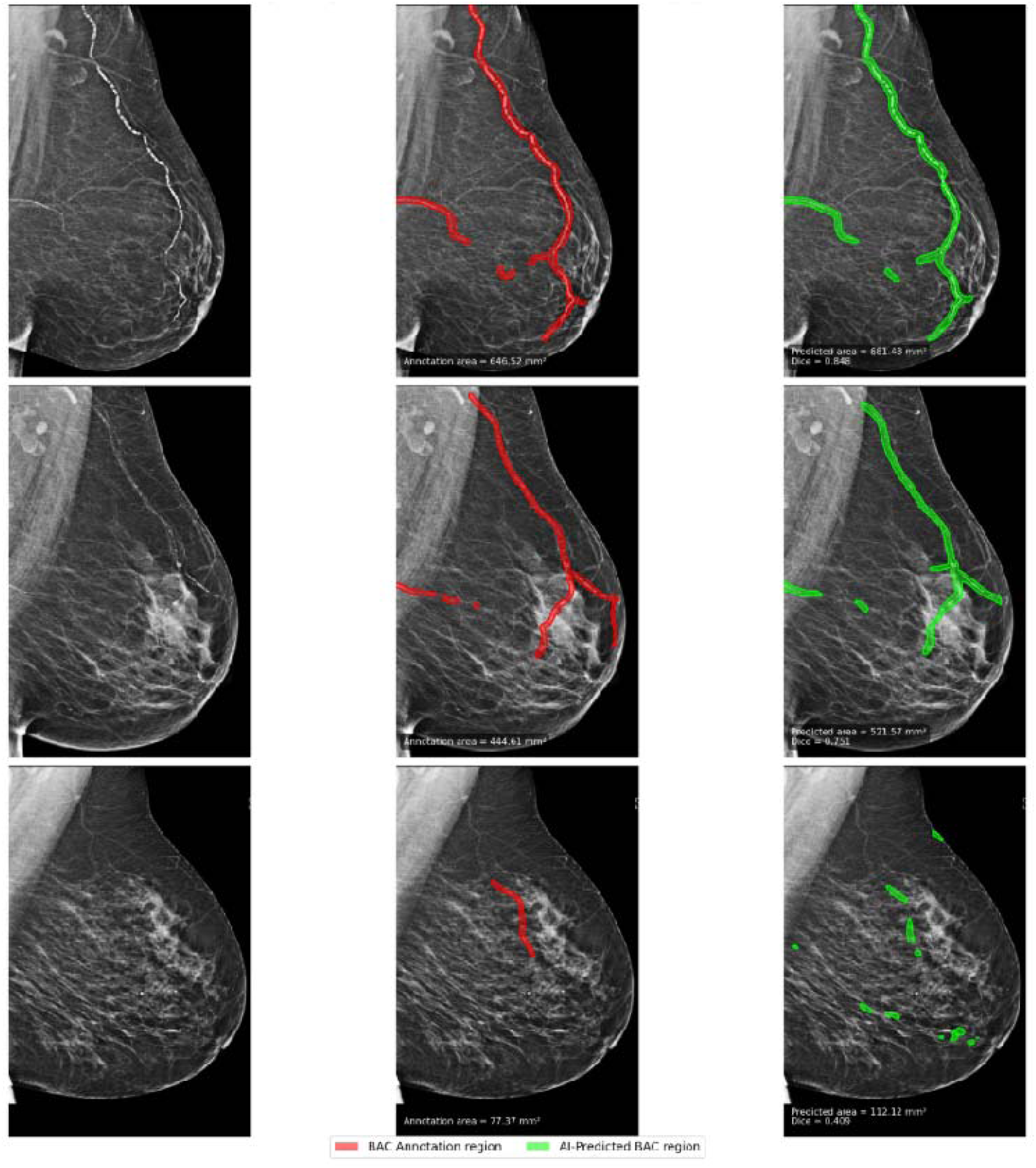
Representative examples of BAC annotation and AI-derived BAC segmentation. Representative screening mammograms showing the original image, manually annotated breas arterial calcification (BAC) regions, and AI-derived BAC regions. The first two rows show examples with close visual agreement between BAC annotation and AI-derived BAC segmentation, whereas the third row shows a failure case with incomplete and false-positive segmentation. Red overlays indicate BAC annotation regions, and green overlays indicate AI-predicted BAC regions. Annotation area, AI-derived area, and Dice similarity coefficient are shown for each example. BAC area is reported in mm².

### Distribution of AI-Derived BAC Burden by Age Group and 10-year MACE Status

AI-derived BAC burden increased with age (**Figure 4**). Among patients without 10-year MACE, median BAC burden was low in younger age groups and increased progressively with age, from 3.76 mm² in patients ≤40 years to 96.94 mm² in patients aged ≥80 years. A similar age-related increase was observed among patients with 10-year MACE, with higher median BAC burden in older age groups, reaching 111.06 mm² in patients aged ≥80 years. Within each age stratum, patients who developed 10-year MACE had higher BAC burden compared with patients without MACE (7 of 10 age strata reached statistical significance at BH-corrected p<0.05). The distribution of BAC burden was highly skewed (skewness=5.67), with a small subset of patients showing substantially higher calcification burden.

**Figure 4.**
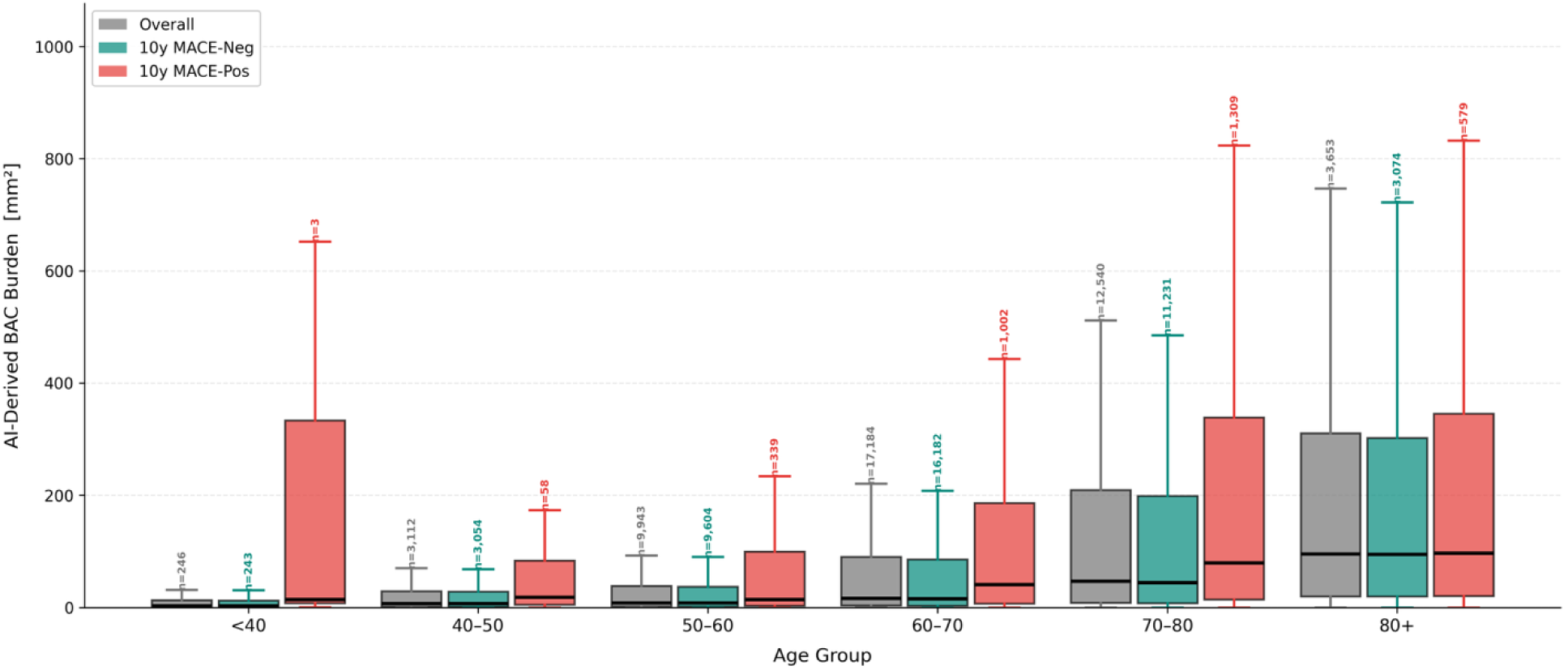
AI-derived BAC burden by age group and MACE status. Box plots show AI-derived breast arterial calcification (BAC) burden among BAC-positive patients, stratified by age group and major adverse cardiovascular event (MACE) status. MACE-positive patients were defined as those who developed MACE within 10 years after the index mammogram; MACE-negative patients did not develop MACE within the 10-year window. Boxes indicate interquartile ranges. Horizontal lines indicate medians. Whiskers indicate minimum-to-maximum range.

### AI-Derived BAC Burden Stratification and MACE Incidence

The BAC burden groups included 155,328 patients with no BAC, 15,560 with low BAC burden (<6.65 mm²), 15,558 with intermediate BAC burden (6.65 to 61.46 mm²), and 15,560 with high BAC burden (>61.46 mm²) (**Figure 5A**). MACE incidence increased across BAC burden categories (**Figure 5B** and **5C**). Five-year MACE incidence was 1.5% among patients with no BAC, 2.8% among patients with low BAC burden, 4.0% among patients with intermediate BAC burden, and 6.9% among patients with high BAC burden (p<0.001). Ten-year MACE incidence was 2.6%, 4.4%, 6.4%, and 10.4% across the same groups, respectively (p<0.001). Kaplan-Meier analysis (**Figure 6**) showed progressively lower MACE-free survival across increasing BAC burden categories; multi-variate log-rank p smaller than 0.0001 for five and ten-year MACE incidence.

**Figure 5.**
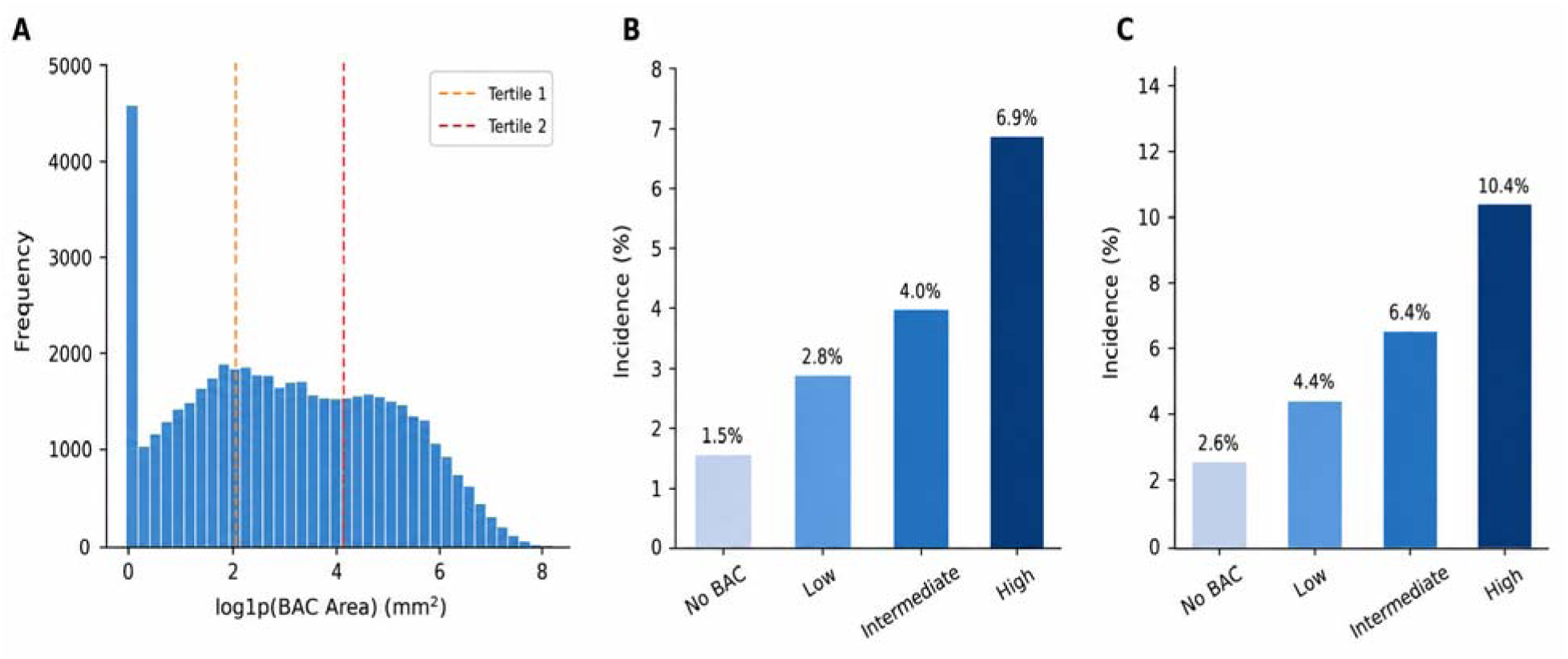
Distribution of AI-derived breast arterial calcification (BAC) burden and MACE incidence across BAC burden categories. (A) Distribution of log1p-transformed AI-derived BAC area among BAC-positive patients. Dashed vertical lines indicate tertile thresholds used to define low, intermediate, and high BAC burden. (B) Five-year incidence of major adverse cardiovascular events (MACE) across BAC burden categories. (C) Ten-year incidence of MACE across BAC burden categories. BAC burden categories were defined as no BAC and tertiles of AI-derived BAC area among BAC-positive patients. P values reflect differences in MACE incidence across BAC burden categories. BAC area is reported in mm².

**Figure 6.**
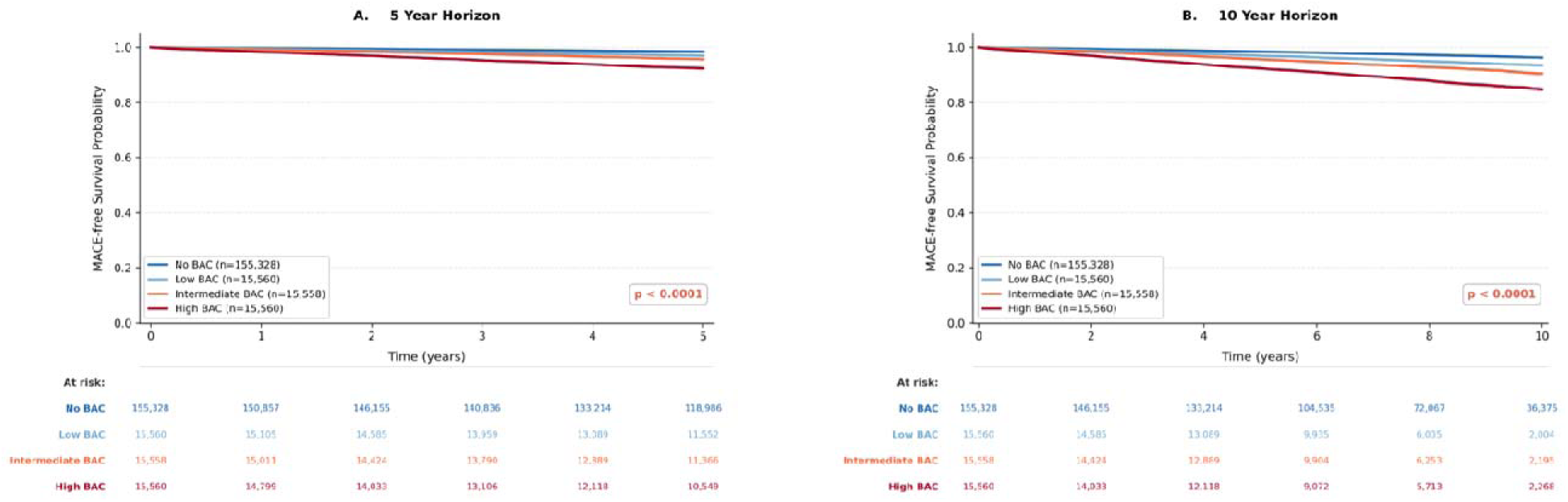
MACE-free survival by AI-derived BAC burden category. Kaplan-Meier curves show MACE-free survival across no, low, intermediate, and high breast arterial calcification (BAC) burden categories over 5-year and 10-year horizons. Low, intermediate, and high BAC categories were defined by tertiles of AI-derived BAC area among BAC-positive patients. P values were calculated using the log-rank test.

### AI-Derived BAC and Prediction of MACE

AI-derived BAC burden was evaluated as a predictor of incident MACE over 5-year and 10-year horizons (**Figure 7**). In univariable analysis, BAC burden alone achieved a 5-year AUROC of 0.6610 [0.644–0.679] and a 10-year AUROC of 0.6498 [0.637–0.663], with a C-index of 0.6545 [0.642–0.668] in the internal test cohort. In the geographically held-out Brooklyn test cohort, BAC burden alone achieved a 5-year AUROC of 0.6579 [0.628–0.686], a 10-year AUROC of 0.6602 [0.635–0.685], and a C-index of 0.6552 [0.630–0.681]. We also evaluated binary BAC (presence vs. absence, 1/0) as a univariable predictor. Binary BAC achieved a 5-year AUROC of 0.6250 [0.611–0.638], a 10-year AUROC of 0.6049 [0.593–0.615], and a C-index of 0.6136 [0.602–0.624] in the internal test cohort, with corresponding values of 0.6274 [0.602–0.651], 0.6234 [0.601–0.645], and 0.6234 [0.601–0.644] in the Brooklyn test cohort.

**Figure 7.**
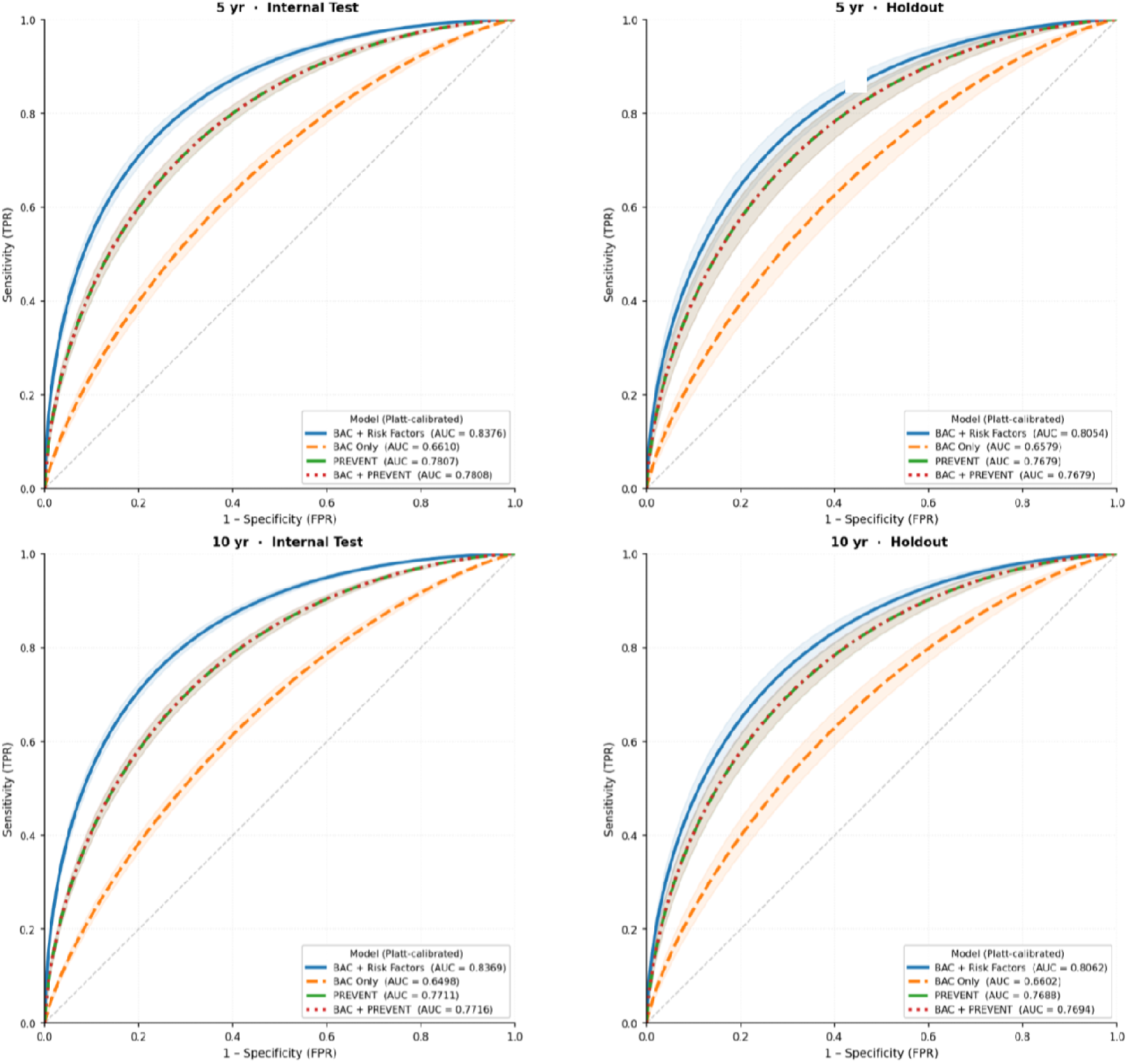
Discrimination performance for MACE prediction across 5-year and 10-year horizons. Receiver operating characteristic curves are shown for calibrated models predicting incident major adverse cardiovascular events (MACE) in the internal test cohort and geographically held-out Brooklyn test cohort. Models included AI-derived breast arterial calcification (BAC) burden alone, PREVENT score alone, BAC plus PREVENT score, and BAC plus clinical risk factors. Shaded regions indicate 95% confidence intervals. AUC indicates area under the receiver operating characteristic curve; FPR, false-positive rate; TPR, true-positive rate.

The PREVENT score achieved a 5-year AUROC of 0.7807 [0.768–0.794], a 10-year AUROC of 0.7711 [0.761–0.782], and a 10-year C-index of 0.7706 [0.760–0.782] in the internal test cohort, and corresponding values of 0.7679 [0.742–0.793], 0.7688 [0.749–0.789], and 0.7634 [0.743–0.785] in the Brooklyn test cohort. Compared with PREVENT alone, the addition of BAC burden to PREVENT score changed the 5-year AUROC from 0.7807 to 0.7808 (+0.0001), the 10-year AUROC from 0.7711 to 0.7716 (+0.0005), and the C-index from 0.7776 / 0.7706 to 0.7777 / 0.7713 for the 5- and 10-year horizons, respectively, in the internal test set. The corresponding changes in the holdout (Brooklyn) test cohort were 0.7679 to 0.7679 (unchanged) for 5-year AUROC, 0.7688 to 0.7694 (+0.0006) for 10-year AUROC, and 0.7662 / 0.7634 to 0.7662 / 0.7640 for the 5- and 10-year C-index, respectively.

## Discussion

In this study of 202,006 women who underwent routine screening mammography, automated deep-learning based quantification of breast arterial calcification (BAC) provided important prognostic information for cardiovascular disease risk. We showed that (1) a deep learning model was able to accurately detect and quantify BAC on full-field digital mammography; (2) AI-detected BAC was present in 23.1% of women, and BAC burden increased with age and was higher among women who subsequently developed major adverse cardiovascular events (MACE); (3) AI-derived BAC burden alone showed modest but reproducible prediction of 5- and 10-year MACE in both the test cohort and the geographically held-out Brooklyn cohort, (4) increasing BAC burden was associated with an increase in MACE incidence and lower MACE-free survival, and (5) adding BAC burden to the PREVENT score produced little improvement in overall discrimination, indicating that BAC should be viewed as a complementary opportunistic imaging biomarker rather than a replacement for established clinical risk assessment.

Our study reinforces the concept of opportunistic imaging, whereby routinely acquired imaging studies are repurposed to extract additional clinically meaningful information without added cost, radiation, or patient burden. More than 50 million mammograms are performed annually in the US, typically beginning in midlife, when cardiovascular disease risk is rising but before many women have come to clinical attention for prevention.^27–29^ Mammography therefore provides a uniquely scalable platform for identifying imaging markers of vascular health in women. Though BAC did not improve risk prediction over the PREVENT score alone in our cohort, it does remain a marker of risk, and making women aware of their cardiovascular disease risk has the potential to motivate healthy behaviors and uptake of interventions such as lipid lowering therapy. The NOTIFY-1 randomized trial found that showing patients coronary artery calcification that was opportunistically detected using an AI algorithm dramatically increased the rate of statin prescription ^30.^

The technical performance of our BAC model supports the feasibility of automated implementation. In the geographically held-out test set, a population with different demographics and clinical characteristics than the development set, our model achieved an image-level BAC detection AUROC of 0.97 and strong agreement with manual BAC area annotation, with a Pearson correlation coefficient of 0.961 overall and 0.950 among BAC-positive cases. Although pixel-level segmentation performance was moderate, the model generated quantitative BAC measures that were sufficiently reliable for downstream population-level risk stratification.

We found a strong association between BAC burden and cardiovascular disease outcomes. Women with no AI-detected BAC had a 5-year MACE incidence of 1.5% and a 10-year incidence of 2.6%, whereas women in the high BAC burden group had corresponding incidences of 6.9% and 10.4%. Intermediate groups showed progressively higher event rates, and Kaplan-Meier analyses demonstrated progressively lower MACE-free survival across increasing BAC categories. The lower performance of binary BAC relative to continuous BAC burden across all cohorts and all-time horizons reflects the inherent loss of granularity when collapsing a continuous measure into a binary classification, as all individuals within the same binary class are assigned an identical predicted risk value regardless of the true extent of calcification. These findings support the biological and clinical relevance of BAC as an imaging biomarker of cumulative vascular injury and suggest that the amount of calcification, not only its presence or absence, is important for risk stratification.

AI-derived BAC burden increased with age and was right-skewed, consistent with vascular calcification as a chronic process that accumulates over time. Importantly, within most age groups, women who developed MACE had higher BAC burden than women who did not develop MACE, suggesting that BAC captures clinically relevant risk information within age groups rather than simply reflecting older age. This is especially relevant for mammography-based prevention because screening often occurs during the transition from lower short-term risk to increasing lifetime cardiovascular risk.

Our findings are consistent with prior studies linking BAC on mammography with cardiovascular outcomes. A recent systematic review and meta-analysis of 45 studies including 68,584 women reported an overall BAC prevalence of 17.1% and found BAC to be associated with traditional risk factors, including diabetes, hypertension, and hyperlipidemia, as well as incident stroke, heart failure, cardiac death, atherosclerotic cardiovascular disease (ASCVD), and all-cause mortality ^31^. Our BAC prevalence of 23.1% is somewhat higher but broadly comparable, likely reflecting differences in the age distribution of the screening population, the use of automated detection, and our definition of BAC burden across standard FFDM views. Importantly, the graded 5- and 10-year event rates observed in our cohort across no, low, intermediate, and high BAC categories provide large-scale support for the conclusion that BAC is not merely an incidental mammographic finding but a marker of systemic vascular risk.

A recent study by Dapamede et al. ^15^ evaluated 123,762 women from two healthcare systems and showed that AI-quantified BAC was associated with MACE and mortality, with mild, moderate, and severe BAC all prognostic after adjustment for PREVENT and each 1-mm² increase in BAC associated with an additional 2%-3% risk of MACE ^15^. Nerlekar et al. studied 21,514 women from the United States and Australia and showed that age-adjusted BAC percentiles predicted MACE beyond ASCVD risk scores. In that study, adding BAC improved the C-statistic from 0.67 to 0.71 and produced an overall net reclassification index of 5% ^32^. Our study extends this literature by evaluating a larger cohort of 202,006 women, using a fully automated deep learning segmentation approach with physical area quantification, and testing model performance in a geographically held-out cohort. At the same time, our finding that adding BAC to PREVENT produced minimal improvement in AUROC or C-index is more conservative than some prior reports. This difference may reflect the improved discrimination provided by the PREVENT score in comparison with earlier risk scores such and the pooled cohort equation as well as differences in cohort composition, outcome definitions, inclusion of mortality, follow-up duration, and BAC scoring methods. Taken together, the literature and our data suggest that the most immediate value of BAC may be not as a replacement for clinical risk models, but as a standardized, visible imaging marker that can prompt clinical risk assessment and preventive care.

Traditional cardiovascular risk models such as PREVENT remain the standard for clinical risk stratification, and our results support that role. PREVENT outperformed BAC burden alone for discrimination of MACE, with 5- and 10-year AUROCs of approximately 0.78 and 0.77 in the internal test cohort, compared with approximately 0.66 and 0.65 for BAC alone. However, discrimination metrics may not fully capture the potential clinical value of BAC, particularly its role as a patient-facing, image-derived marker that could prompt risk discussion, lipid assessment, PREVENT calculation, and preventive treatment when indicated.

The integration of deep learning-enabled BAC quantification into routine mammography workflows has several potential clinical implications. It enables large-scale cardiovascular risk screening in asymptomatic women without requiring additional imaging or clinician-initiated testing. In addition, it may help activate prevention in women whose cardiovascular risk is under-recognized or whose clinical risk assessment is incomplete because laboratory or risk-factor data are unavailable. Automated quantification allows for standardized and reproducible reporting of BAC burden, which could be incorporated into structured mammography reports and linked to clinical decision support. A pragmatic implementation pathway would report BAC in a standardized manner and connect the result to PREVENT score calculation, lipid evaluation, primary care or cardiology follow-up, and evidence-based prevention when appropriate.

Our study has several limitations. This was a retrospective study conducted within a single health system, which may limit generalizability. However, we validated the algorithm in a cohort with different demographic and clinical characteristics, and the algorithm showed similar performance. Cardiovascular outcomes were ascertained using diagnosis and procedure codes in the electronic health record, which may introduce misclassification and may miss events occurring outside the health system. Cause-specific cardiovascular mortality was not available and therefore was not included in the MACE definition. We did not incorporate use of medications such as lipid lowering therapy in our risk modeling.

In conclusion, deep learning-based quantification of breast arterial calcifications from routine screening mammograms is feasible and provides clinically meaningful prognostic information for cardiovascular risk assessment in women. BAC burden was associated with age and subsequent MACE. Although BAC did not significantly improve discrimination when added to PREVENT, its automated detection on a widely performed screening examination may provide an important opportunity to make cardiovascular disease risk visible and actionable. These findings support further prospective evaluation of BAC reporting and clinical decision support as a scalable strategy for cardiovascular prevention in women.

## Data Availability

All data generated or analyzed during the study are included in the published paper.

